# Targeted sequencing of Parkinson’s disease loci genes highlights *SYT11, FGF20* and other associations

**DOI:** 10.1101/2020.05.29.20116111

**Authors:** Uladzislau Rudakou, Eric Yu, Lynne Krohn, Jennifer A. Ruskey, Farnaz Asayesh, Yves Dauvilliers, Dan Spiegelman, Lior Greenbaum, Stanley Fahn, Cheryl H. Waters, Nicolas Dupré, Guy A. Rouleau, Sharon Hassin-Baer, Edward A. Fon, Roy N. Alcalay, Ziv Gan-Or

**Affiliations:** Department of Human Genetics, McGill University, Montréal, QC, H3A 1A1, Canada; Montreal Neurological Institute, McGill University, Montréal, QC, H3A 1A1, Canada; Department of Neurology and Neurosurgery, McGill University, Montréal, QC, H3A 1A1, Canada; National Reference Center for Narcolepsy, Sleep Unit, Department of Neurology, Gui-de-Chauliac Hospital, CHU Montpellier, University of Montpellier, Inserm U1061, Montpellier, France; The Danek Gertner Institute of Human Genetics, Sheba Medical Center, Tel Hashomer, Ramat Gan, Israel; The Joseph Sagol Neuroscience Center, Sheba Medical Center, Tel Hashomer, Ramat Gan, Israel; Sackler Faculty of Medicine, Tel Aviv University, Tel Aviv, Israel; Department of Neurology, College of Physicians and Surgeons, Columbia University Medical Center, New York, NY 10032, USA; Division of Neurosciences, CHU de Québec, Université Laval, Québec City, QC, G1V 0A6, Canada; Department of Medicine, Faculty of Medicine, Université Laval, Québec City, QC, G1V 0A6, Canada; Department of Neurology, Sheba Medical Center, Tel Hashomer, Ramat Gan, Israel; Movement Disorders Institute, Sheba Medical Center, Tel Hashomer, Ramat Gan, Israel; Taub Institute for Research on Alzheimer’s Disease and the Aging Brain, College of Physicians and Surgeons, Columbia University Medical Center, New York, NY 10032, USA

**Keywords:** Parkinson’s disease, genetic fine mapping, risk loci, association study, sequencing

## Abstract

Genome-wide association studies (GWAS) have identified numerous loci associated with Parkinson’s disease. The specific genes and variants that drive the associations within the vast majority of these loci are unknown. We aimed to perform a comprehensive analysis of selected genes to determine the potential role of rare and common genetic variants within these loci. We fully sequenced 32 genes from 25 loci previously associated with Parkinson’s disease in 2,657 patients and 3,647 controls from three cohorts. Capture was done using molecular inversion probes targeting the exons, exon-intron boundaries and untranslated regions (UTRs) of the genes of interest, followed by sequencing. Quality control was performed to include only high-quality variants. We examined the role of rare variants (minor allele frequency < 0.01) using optimized sequence Kernel association tests (SKAT-O). The association of common variants was estimated using regression models adjusted for age, sex and ethnicity as required in each cohort, followed by a meta-analysis. After Bonferroni correction, we identified a burden of rare variants in *SYT11, FGF20* and *GCH1* associated with Parkinson’s disease. Nominal associations were identified in 21 additional genes. Previous reports suggested that the *SYT11* GWAS association is driven by variants in the nearby *GBA* gene. However, the association of *SYT11* was mainly driven by a rare 3’ UTR variant (rs945006601) and was independent of *GBA* variants (p=5.23E-05 after exclusion of all *GBA* variant carriers). The association of *FGF20* was driven by a rare 5’ UTR variant (rs1034608171) located in the promoter region. The previously reported association of *GCH1* with Parkinson’s Disease is driven by rare nonsynonymous variants, some of which are known to cause dopamine-responsive dystonia. We also identified two *LRRK2* variants, p.Arg793Met and p.Gln1353Lys, in ten and eight controls, respectively, but not in patients. We identified common variants associated with Parkinson’s disease in *MAPT, TMEM175, BST1*, *SNCA* and *GPNMB* which are all in strong linkage disequilibrium (LD) with known GWAS hits in their respective loci. A common coding *PM20D1* variant, p.Ile149Val, was nominally associated with reduced risk of Parkinson’s disease (OR 0.73, 95% CI 0.60-0.89, p=1.161E-03). This variant is not in LD with the top GWAS hits within this locus and may represent a novel association. These results further demonstrate the importance of fine mapping of GWAS loci, and suggest that *SYT11, FGF20*, and potentially *PM20D1, BST1* and *GPNMB* should be considered for future studies as possible Parkinson’s disease-related genes.

## Introduction

Genome-wide association studies (GWASs) are an important tool for identifying genetic associations with complex disorders such as Parkinson’s Disease (Nalls *et al*., 2014; Chang *et al*., 2017; Nalls *et al*., 2019). GWASs examine the association of multiple common variants across the genome with specific traits. Typically, each associated locus includes multiple genes, and very often GWAS alone cannot identify the specific gene or variant within each locus that drives the association. Furthermore, GWASs generally overlook rare variants, while accumulation of rare variants on a specific common allele may drive some of these associations. Even if not located on a specific common allele, rare variants in GWAS genes may contribute to disease. For example, rare variants in GWAS loci genes including *SNCA, GBA, LRRK2, VPS13C* and *GCH1* may cause Mendelian parkinsonism or strongly increase the risk of Parkinson’s disease. (Polymeropoulos *et al*., 1997; Sidransky *et al*., 2009; Ran *et al*., 2016; Emelyanov *et al*., 2018; Amaral *et al*., 2019; Paisan-Ruiz *et al*., 2004; Khan *et al*., 2005; Lesage *et al*., 2016; Jansen *et al*., 2017; Darvish *et al*., 2018; Schormair *et al*., 2018; Rudakou *et al*., 2020; Mencacci *et al*., 2014; Guella *et al*., 2015; Lewthwaite *et al*., 2015; Xu *et al*., 2017; Yoshino *et al*., 2018; Rudakou *et al*., 2019).

Recently, a large-scale GWAS has identified 78 loci associated with risk of Parkinson’s disease (Nalls *et al*., 2019), yet only for a few of them we have identified the specific genes and variants that drive the association. In some of these loci, there are multiple hits, suggesting that more than one variant, or perhaps more than one gene within the locus, can be associated with Parkinson’s disease. Fine-mapping of GWAS loci may help identifying these genes and variants, as was done for *SNCA* (Soldner *et al*., 2016; Krohn *et al*., 2019b), *VPS13C* (Lesage *et al*., 2016; Jansen *et al*., 2017; Darvish *et al*., 2018; Schormair *et al*., 2018; Rudakou *et al*., 2020) and *TMEM175* (Krohn *et al*., 2020). In the *SNCA* region, fine mapping has identified independent 5’ and 3’ variants that drive the associations with different synucleinopathies (Krohn *et al*., 2019b). Similarly, a potentially protective haplotype in *VPS13C* (Rudakou *et al*., 2020) and common nonsynonymous variants in *TMEM175* have been associated with Parkinson’s Disease through fine mapping, and in the case of *TMEM175* also by functional studies (Jinn *et al*., 2019; Krohn *et al*., 2020).

In the current study, we fully sequenced 32 genes from 25 different Parkinson’s disease GWAS loci in a total of 2,657 cases and 3,647 controls. We examined the association of common and rare variants with Parkinson’s Disease, including the role of rare bi-allelic variants.

### Materials and Methods

### Study Population

The study population included, consecutively recruited, unrelated patients with Parkinson’s Disease (n=2,657) and controls (n=3,647) from three cohorts, collected at McGill University (Quebec, Canada and Montpellier, France), Columbia University (New York, NY, USA), and the Sheba Medical Center (Israel). The McGill cohort was recruited in Quebec, Canada (in part with the assistance of the Quebec Parkinson’s Network (Gan-Or *et al*., 2020)) and in Montpellier, France. It includes 1,026 Parkinson’s Disease patients (mean age 61.5±11.3, 62.6% male) and 2,588 controls (mean age 54.5±14.2, 47.4% male), all unrelated of French-Canadian/French ancestry. The Columbia cohort includes 1,026 Parkinson’s Disease patients (mean age 59.4±11.7, 64.2% male) and 525 controls (mean age 64.2±10.1, 37.1% male). This cohort has been previously described in detail, and is mainly composed of participants with European descent, but 21.2% of Parkinson’s Disease patients and 18.7% controls are of Ashkenazi Jewish (AJ) origin (Alcalay *et al*., 2016). The Sheba cohort included 605 Parkinson’s Disease patients (mean age 60.7±11.9, 62.3% male) and 534 controls (mean age 34.0±7.0, 55.8% male), all of full AJ origin; previously described in more details (Ruskey *et al*., 2019). Movement disorder specialists diagnosed Parkinson’s Disease with UK brain bank criteria (Hughes *et al*., 1992) or the MDS clinical diagnostic criteria (Postuma *et al*., 2015).

As detailed in the Statistical Analyses subsection, due to the differences in age and sex, statistical analyses were adjusted and included age and sex as co-variates. To account for ethnical heterogeneity of the Columbia cohort, an ethnicity covariate was also introduced for the analysis of this cohort (GWAS data was not available, therefore the reported ethnicity was used and not principal components). All three cohorts were sequenced in the same lab (McGill University), using the same protocol.

### Standard Protocol Approvals, Registrations, and Patient Consents

The institutional review boards approved the study protocols and informed consent was obtained from all individual participants before entering the study.

### Target genes

A recent Parkinson’s Disease GWAS identified 78 loci, however the current study was designed and performed before its publication and the target genes were taken from earlier literature (Nalls *et al*., 2014; Chang *et al*., 2017). Our target genes included: *ACMSD, BST1, CCDC62, DDRGK1, FGF20, GAK, GCH1, GPNMB, HIP1R, INPP5F, ITGA8, LAMP3, LRRK2, MAPT, MCCC1, PM20D1, RAB25, RAB29, RIT2, SCARB2, SETD1A, SIPA1L2, SLC41A1, SNCA, SREBF1, STK39, STX1B, SYT11, TMEM163, TMEM175, USP25* and *VPS13C*. These 32 genes included three known Parkinson’s disease related genes *(LRRK2, GCH1, SNCA)* found in GWAS loci in order to perform full sequencing assessment. The other 29 genes were selected for sequencing based on assessment that included effects of quantitative trait loci, brain expression, potential association with known Parkinson’s disease-causing genes and involvement in pathways potentially involved in Parkinson’s disease, such the autophagy-lysosomal pathway, mitochondria quality control and endolysosomal recycling.

### Sequencing

The coding regions, exon-intron boundaries and the 5’ and 3’ untranslated regions (UTRs) of the genes listed above were targeted using molecular inversion probes (MIPs), as previously described (O’Roak *et al*., 2012). All MIPs used to sequence the genes of the study are detailed in Supplementary Table 1. Targeted DNA capture and amplification was done as previously described (Ross *et al*., 2016), and the full protocol is available upon request. The library was sequenced using Illumina HiSeq 2500/4000 platform at the McGill University and Genome Quebec Innovation Centre. Sanger sequencing was performed for rare variants with significant association to Parkinson’s disease. Primers and conditions are available upon request.

**Table 1:**
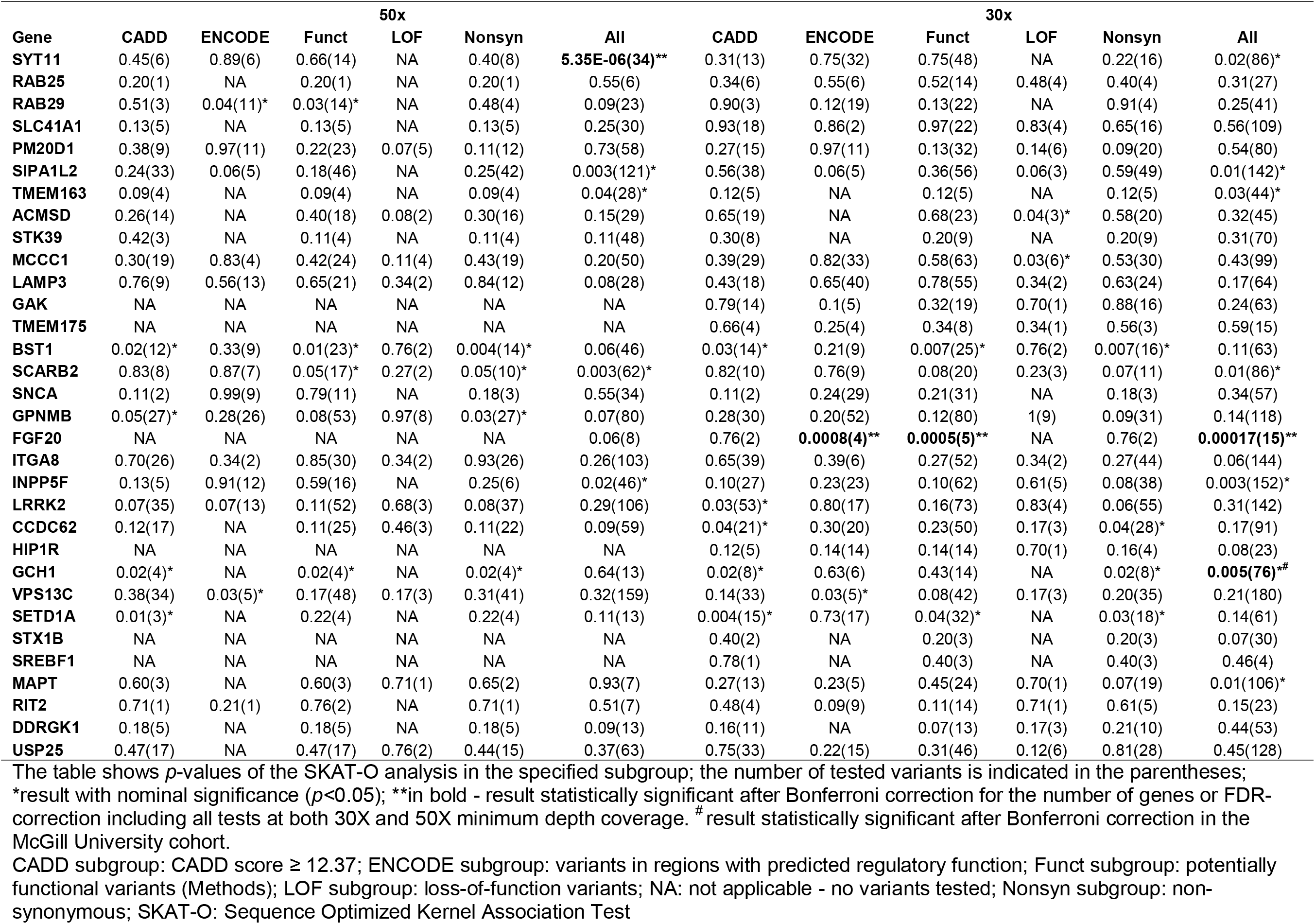
Results of SKAT-O analysis in the merged cohorts at 30x and 50x minimum depth of coverage.

### Data processing and quality control

Reads were mapped to the human reference genome (hg19) with Burrows-Wheeler Aligner (Li and Durbin, 2009). Post-alignment quality control and variant calling was performed using Genome Analysis Toolkit (GATK, v3.8) (McKenna *et al*., 2010). Variant annotation was done with ANNOVAR (Wang *et al*., 2010). Data on the frequency of each variant in different populations were extracted from the public database Genome Aggregation Database (GnomAD) (Lek *et al*., 2016).

The quality control pipeline that was used in this study is available on GitHub (https://github.com/gan-orlab/MIPVar.git). All calls with less than 25% of the reads containing the observed allele have been removed. The remaining quality control (QC) was performed using GATK and PLINK software v1.9 (Purcell *et al*., 2007). Genotyping quality (GQ) < 30 was used as a threshold for variant exclusion. Different depths of coverage thresholds were used for common and rare variants. Common variants threshold for inclusion was ≥ 15x, while rare variants (minor allele frequency [MAF] < 0.01) thresholds were more stringent and repeated twice, at ≥ 30x and ≥ 50x. Variants with genotype rate lower than 90% were excluded. Variants that deviated from Hardy-Weinberg equilibrium set at *p ≤* 0.001 were filtered out. Threshold for missingness difference between cases and controls was set at *p =* 0.05 and the filtration script adjusted it with Bonferroni correction. Samples with average genotyping rate of less than 90% were excluded.

### Statistical analysis

The burden of rare variants in each of the target genes was tested using the optimized sequence Kernel association test (SKAT-O, R package) (Lee *et al*., 2012). In addition, we examined the burden of specific variant subgroups including: i) Funct.: Potentially functional included all nonsynonymous mutations, stop gain/loss, frameshift mutations, splicing variants located within two base pairs of exon-intron junctions, and variants that were flagged by ENCODE (explained below). ii) NS: nonsynonymous variants. iii) LOF: loss-of-function variants included stop gain/loss, frameshift, and splicing variants located within two base pairs of exon-intron junctions. iv) CADD: variants with the Combined Annotation Dependent Depletion (CADD) score ≥ 12.37; this threshold indicates the top 2% variants predicted to be the most damaging in the genome (Amendola *et al*., 2015). v) ENCODE: variants that are predicted to have regulatory activity, such as promoter, enhancer, activator and repressor, based on the chromatin profiling studies (Ernst *et al*., 2011). To increase power, rare variants were assessed in the three merged cohorts (since rare variants are less affected by ethnicity). Before merging, allele frequencies were compared between the control groups and between the patient groups. We have also compared allele frequencies between young controls and elderly controls specifically. Since there were no significant differences when comparing the control groups and the patient groups, we could merge them for the analysis. Analysis of the individual cohorts was also performed post-hoc. Similarly to exome-wide burden analyses of rare variants (Kenna *et al*., 2016), Bonferroni correction for statistical significance was performed based on the number of genes that were tested, and the thresholds are *p* < 1.85E-03 for genes with variant coverage of ≥ 50x and *p* < 1.56E-03 at ≥ 30x. Since the different categories and coverages are not independent, we have also applied false discovery rate (FDR) correction to account for all analyses in all categories combined. To determine if an identified association was mainly driven by a single variant, we performed a ‘leave-one-out’ analysis, in which we systematically excluded a single variant each time for a certain gene and examine whether excluding this variant affected the observed results. Variants that their exclusion led to non-significant or to a highly reduced statistical significance were considered as main drivers of the initially observed association.

The association between common variants and Parkinson’s Disease was examined by logistic regression models using PLINK v1.9, with the status (patient or control) as a dependent variable, age and sex as covariates in all cohorts. Ethnicity as an additional covariate in the Columbia University cohort due to the ethnical heterogeneity. Meta-analysis of our three cohorts was conducted using METAL (R package) (Willer *et al*., 2010) with a fixed effect model. Cochran’s Q-test was applied to test for residual heterogeneity. LD of common variants and their respective GWAS top marker was examined using the LDlink online tool LDmatrix (https://ldlink.nci.nih.gov/?tab=ldmatrix) (Machiela and Chanock, 2015). Effects of common and rare variants on expression levels (eQTL) or splicing (sQTL) was evaluated using GTEx Portal online tool (https://www.gtexportal.org/home/).

### Data availability

Anonymized data is available upon request by any qualified investigator.

## Results

### Coverage and variants identified

The average coverage for all 32 genes included in the current analysis was 642X, with a range of 86-1,064 (median 688) across all genes. An average of 97% of the targeted regions was covered at ≥ 15X, 96% at ≥ 30X, and 92% at ≥ 50X. Supplementary Table 2 details the average coverage and the percent coverage for each gene. At coverage of ≥ 30X, we found 3,327 rare variants and at ≥ 50X, 1,713 rare were identified (Supplementary Tables 3 and 4). A total of 311 common variants were identified across all genes in all three cohorts (Supplementary Table 5) with a coverage of ≥15X.

**Table 2.**
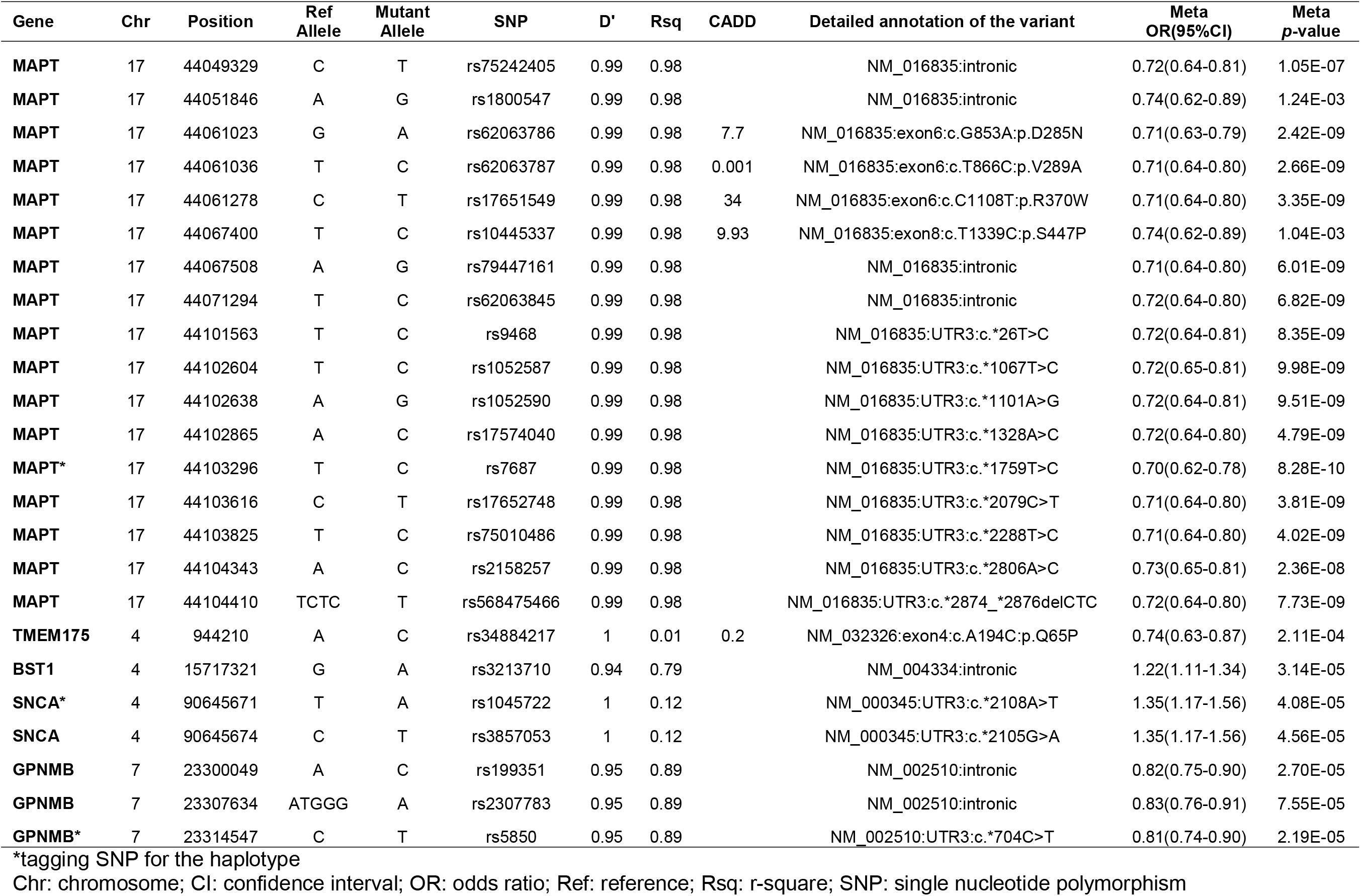
The common variants with statistically significant association with Parkinson’s Disease in the meta-analysis of three cohorts.

### Rare variants in *SYT11, FGF20* and *GCH1* are associated with Parkinson’s disease

Table 1 details the SKAT-O results of the analysis in the three combined cohorts, and Supplementary Table 6 details the results per cohort. After Bonferroni correction, *SYT11, FGF20* and *GCH1* were associated with Parkinson’s Disease. Nominal associations were identified in 21 genes (Table 1, Supplementary Table 6). When considering all variants with MAF < 0.01 in *SYT11*, there was a strong association with Parkinson’s Disease *(p* = 5.3E-06, Table 1, FDR-corrected p=0.0017, Supplementary Table 7), mainly driven by a *SYT11* 3’ UTR variant (rs945006601) with MAF of 0.004 in Parkinson’s Disease patients and 0.00015 in controls (OR = 27.0, 95%CI 3.6-202.6, *p* = 0.0013). This association was identified at a coverage of ≥ 50X, which provides high certainty that rare variants are being called correctly. We further sequenced this variant using Sanger sequencing, and all carriers have been confirmed, as well as randomly selected non-carriers. At 30X, the association was weaker (p=0.02, Table 1), possibly due to inclusion of other variants with coverage of between 30-50X that diluted the association. It was previously reported that variants in the *SYT11* locus are in LD with known *GBA* variants such as p.Glu326Lys and p.Asn370Ser, and that the association seen with *SYT11* is driven by the *GBA* Variants (Blauwendraat *et al*., 2018). However, the rs945006601 *SYT11* variant that drives the association in the current study is not in LD with these variants, but with the wild-type *GBA* allele. To further test if the identified association in *SYT11* is driven by other *GBA* variants, we removed carriers of all *GBA* variants that were ever reported to be potentially pathogenic, and the association of *SYT11* with PD remained statistically significant (*p*=5.23E-05).

When analyzing *FGF20*, a burden of all rare variants was demonstrated (p=0.0002, Table 1, FDR-corrected p=0.026), mainly driven by a rare 5’ UTR variant, rs1034608171, which is located in the promoter region of the gene, with MAF of 0.002 in Parkinson’s Disease patients and 0.00015 in controls (OR = 13.4, 95% CI 1.7-106.2, *p* = 0.014). This association was discovered at a coverage of ≥ 30X, but at ≥ 50X it was not identified since the average coverage of this variant was below 50X. However, the variant was confirmed using Sanger sequencing.

The association of *GCH1* with Parkinson’s Disease is driven by rare nonsynonymous variants (Supplementary Tables 3 and 4), some of which are known to cause dopamine responsive dystonia as we have previously reported (Rudakou *et al*., 2019). We repeated the analyses above focusing only on early onset PD patients with age at onset <50 years (n=517). We did not identify any associations at coverages of ≥ 30X or ≥ 50X (Supplementary Tables 8 and 9, respectively), yet this is an underpowered analysis, and studies in larger cohorts of early onset PD are necessary.

After removing the *LRRK2* p.Gly2019Ser variant from the analysis (a total of 179 carriers, 10 carriers form the McGill cohort, 82 from the Columbia cohort and 87 in Sheba cohort), there was no statistically significant burden of other rare *LRRK2* variants in our cohorts. However, we did identify two variants, p.Arg793Met (rs35173587) and p.Gln1353Lys (rs200526782), in ten and eight controls, respectively, but not in Parkinson’s Disease. Neither of these two variants had a statistically significant association after Bonferroni correction.

### Association of common variants with potential functional effects in known Parkinson’s disease loci

To examine the association of common variants in the 32 genes, we performed logistic regression adjusted for age and sex in all three cohorts, and also adjusted for ethnicity in the Columbia University cohort. Additionally, our three cohorts were meta-analyzed. After Bonferroni correction we found 24 variants from five genes associated with risk of Parkinson’s Disease (Table 2). Forest plots in Fig. 1 show the meta-analysis results for the tagging variants of these five genes. We further examined the LD of these variants with the known Parkinson’s Disease-associated variants from the most recent Parkinson’s Disease GWAS, and their potential functional effects (detailed in Methods) in ENCODE, GTEx and UCSC genome browser. In the *MAPT* locus, 17 common variants at full LD, all represent the *MAPT* H2 haplotype, were associated with reduced risk of Parkinson’s Disease (Table 2). The variants identified in *TMEM175, BST1, SNCA* and *GPNMB* are all in strong LD with known GWAS hits in their respective loci (Table 2). Two *SNCA* 3’ UTR variants were in full LD between themselves, and in strong LD with rs356219, a known 3’ GWAS hit in the *SNCA* locus. These two variants are located within a strong enhancer region and are associated with altered splicing of *SNCA* in the

**Figure 1.**
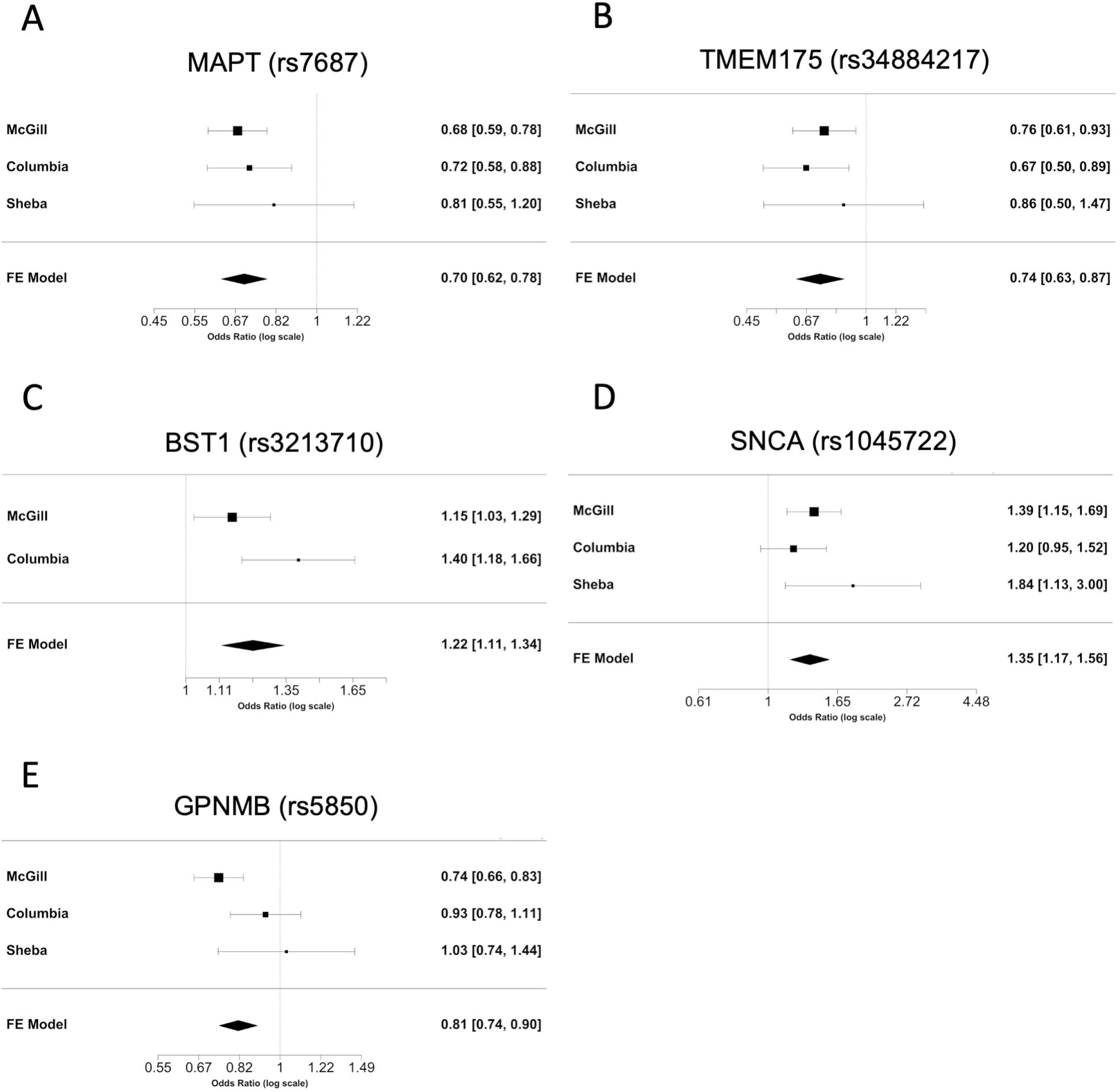
Forest plots, meta-analyses of common variants in the three cohorts. Each panel presents the individual odds ratio and 95% confidence interval for each cohort, and the results of the meta-analysis for the tagging variant of six loci identified in our study. Meta-analysis *p-*values: A. *MAPT* (*p=*8.28E-10); B. *TMEM175* (*p=*2.11E-04); C. *BST1* (*p=*3.14E-05); D. *SNCA* (*p=*4.08E-05); E. *GPNMB* (*p=*2.19E-05)

Cortex (Fig. 2). Three *GPNMB* variants were identified, one of which (rs5850) is located at the 3’ UTR of *GPNMB* and associated with reduced expression of *GPNMB* in numerous brain regions including the substantia nigra, putamen and cortex (Supplementary Fig. 1). A coding variant in *PM20D1*, p.Ile149Val (rs1891460), was nominally associated with reduced risk of Parkinson’s Disease (OR = 0.73, 95% CI 0.60-0.89, *p* = 1.161E-03, specific frequencies in each cohort are in Supplementary Table 5). This variant is located within the *PARK16* region which also includes *RAB29* (previously named *RAB7L1), SLC41A1* and other genes. However, this variant is not in LD with the top GWAS hits within this locus (r^2^ = 0.04, D’ = 0.46).

**Figure 2.**
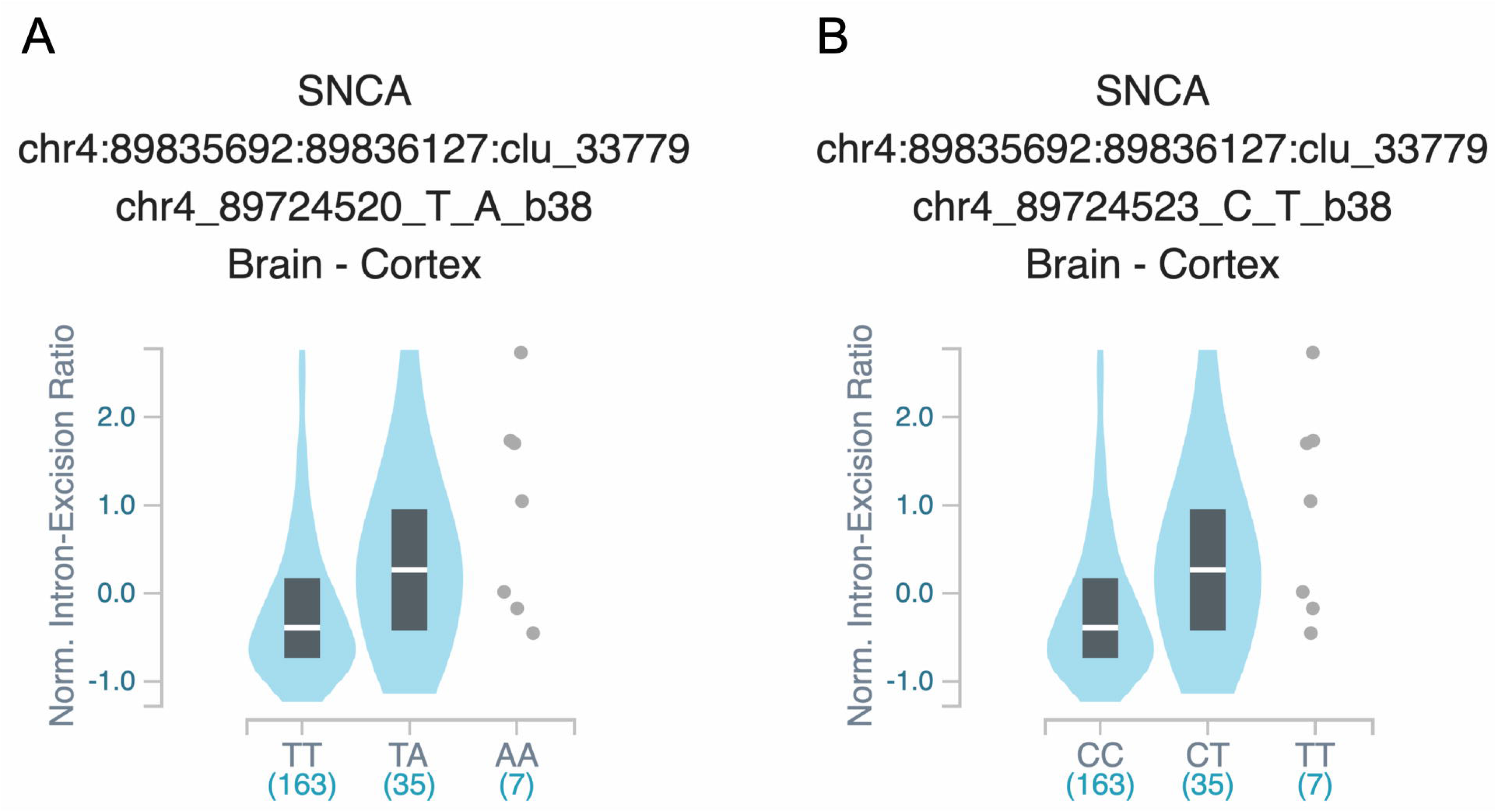
Effect of 3’UTR *SNCA* variants on splicing. Figure is processed from GTeX (https://www.gtexportal.org), and represent the splicing quantitative trait locus (sQTL) effects of: **A**. rs1045722 in the 3’ UTR of *SNCA* and **B**. rs3857053 in the 3’ UTR of *SNCA*

## Discussion

In the current study, we examined the role of common and rare variants in 32 genes located within known Parkinson’s Disease risk loci, by performing targeted next generation sequencing of the coding and 5’ and 3’ untranslated regions, followed by association tests. We found a statistically significant burden, after Bonferroni correction, of rare variants in *SYT11, FGF20* and *GCH1*, and nominal associations (*p<*0.05) with other genes and variants (Table 1, Supplementary Tables 3,4,6). In addition, we have identified common coding and putative regulatory variants associated with Parkinson’s Disease in *MAPT, TMEM175, BST1, SNCA* and *GPNMB*, including a potential novel association with a nonsynonymous variant in *PM20D1*, p.Ile149Val, which is nominally significant and requires further confirmation.

Three genes, *SYT11*, *FGF20* and *GCH1* were identified with a statistically significant burden of rare variants comparing Parkinson’s Disease to controls. *SYT11* is located close to the *GBA* locus, and it was reported that the GWAS signal from *SYT11-GBA* locus in Parkinson’s Disease can be attributed to coding *GBA* variants (Blauwendraat *et al*., 2018). The *SYT11* variant mainly responsible for the association in our data is in weak LD with two coding *GBA* variants, p.Glu326Lys (p.E326K) and p.Asn370Ser (p.N370S). After exclusion of carriers of these variants, and further exclusion of all other *GBA* variants, the association of *SYT11* remained strong. These results suggest that *SYT11* may independently be associated with risk of PD. The Protein encoded by *SYT11*, Synaptotagmin 11, belongs to the synaptotagmin family, known calcium sensors mediating calcium-dependent regulation of membrane trafficking in synaptic transmission (Xie *et al*., 2017). Synaptotagmin 11 is involved in regulation of endocytosis and vesicle recycling process, which is essential for neurotransmission (Wang *et al*., 2016; Xie *et al*., 2017). Overexpression of Synaptotagmin 11 in mice led to impairment of DA transmission (Wang *et al*., 2018). Synaptotagmin 11 was also reported to be a physiological substrate for E3 ubiquitin ligase – parkin, playing a critical role in parkin-related neurotoxicity (Wang *et al*., 2018). Involvement of Synaptotagmin 11 in other mechanisms potentially related to Parkinson’s Disease such as autophagy-lysosomal pathway (Bento *et al*., 2016), phagocytosis of α-synuclein fibrils (Du *et al*., 2017), and repair of injured astrocytes via lysosome exocytosis regulation (Sreetama *et al*., 2016), suggests that *SYT11* needs to be considered as a target for further research in PD. Fibroblast growth factor 20 (*FGF20*) encodes a neurotrophic factor preferentially expressed in the substantia nigra pars compacta. It regulates central nervous system development and function (International Parkinson Disease Genomics Consortium, 2011) and plays a role in dopaminergic neurons differentiation and survival (Itoh and Ohta, 2013). There is a number of conflicting reports about FGF20 overexpression increasing α-synuclein levels in dopaminergic neurons (Wang *et al*., 2008; Wider *et al*., 2009; Sekiyama *et al*., 2014; Tarazi *et al*., 2014). The rare variant (rs1034608171) that drives the association in our data is located in a regulatory region and its potential functional effects would require additional studies. *GCH1* encodes GTP-cyclohydrolase 1, an essential enzyme for the dopamine synthesis pathway in the nigrostriatal cells (Kurian *et al*., 2011). Rare *GCH1* variants may cause DOPA-responsive dystonia, and the same variants have also been associated with Parkinson’s Disease (Mencacci *et al*., 2014; Rudakou *et al*., 2019). Since these mutations are rare, they only reached statistical significance in the McGill cohort, as we have previously described (Rudakou *et al*., 2019). The two *LRRK2* variants, p.Arg793Met and p.Gln1353Lys, that were found only in controls require additional studies to examine their potential protective roles in Parkinson’s Disease. *LRRK2* is a well-validated Parkinson’s Disease related gene (Bandres-Ciga *et al*., 2020).

All 17 *MAPT* variants that were found to be significantly associated with Parkinson’s Disease are in perfect LD with each other, and represent the H2 haplotype, known to be associated with reduced risk of Parkinson’s Disease (Li *et al*., 2018). The identified coding variants in *MAPT* are located within exons that are likely not expressed in the CNS (Caillet-Boudin *et al*., 2015), suggesting that the association in this locus may be due to regulatory variants. *TMEM175* is located in the fourth most significant GWAS locus in Parkinson’s Disease (Nalls *et al*., 2019), and encodes a transmembrane potassium channel responsible for K^+^ conductance in endosomes and lysosomes (Cang *et al*., 2015). TMEM175 regulates lysosomal function through maintenance of lysosomal membrane potential and luminal pH stability, as well as its role in autophagosome-lysosome fusion (Lee *et al*., 2017). In a recent genetic and functional study, two coding *TMEM175* variants, including the variant identified here, were shown to be associated with risk of Parkinson’s Disease, risk of REM-sleep behavior disorder, and glucocerebrosidase activity (Krohn *et al*., 2019a).

The *BST1* and *SNCA* variants associated with risk of Parkinson’s Disease in the current study are in strong LD with the known GWAS markers in their respective loci. The *BST1* variant is intronic without known function, and the *SNCA* 3 ’UTR variants are located within strong *SNCA* enhancers. Whether these *SNCA* variants drive the association with Parkinson’s Disease and their potential functional effects require additional studies. *GPNMB* encodes the glycoprotein nonmetastatic melanoma protein B a type 1, which is a transmembrane glycoprotein shown to have a role in melanoma (Taya and Hammes, 2018). Patients with melanoma have an increased risk of Parkinson’s Disease (Liu *et al*., 2011), and Parkinson’s Disease patients have an increased risk for melanoma (Bertoni *et al*., 2010). GPNMB may have a role in the lysosome (Tomihari *et al*., 2009) and in regulation of microglial inflammation (Dzamko *et al*., 2015; Herrero *et al*., 2015). While the expression of GPNMB in brain is overall low, the two variants identified in the current study are associated with reduced GPNMB expression in various brain tissues (Supplementary Fig. 1). One of the variants, rs5850, is located within the 3’ UTR of *GPNMB*, but there is no clear regulatory function for this variant.

Peptidase M20 Domain Containing 1 (*PM20D1*) encodes a protein that regulates the production of N-fatty-acyl amino acids which have a function of chemical uncouplers of mitochondrial respiration (Long *et al*., 2016). Mitochondrial quality control is likely important in Parkinson’s Disease, and genes involved in mitophagy are involved in familial forms of Parkinson’s Disease (Park *et al*., 2018). *PM20D1* is located within the *PARK16* locus which also contains *SLC45A3, NUCKS1, RAB29* (previously named *RAB7L1*) and *SLC41A1* (Satake *et al*., 2009; Simon-Sanchez *et al*., 2009; Nalls *et al*., 2019).Several studies have highlighted *RAB29* as the potential gene associated with Parkinson’s Disease in this locus (Gan-Or *et al*., 2012),as it was shown to interact with *LRRK2* (MacLeod *et al*., 2013) and affect lysosomal function (Eguchi *et al*., 2018). In the current study we identified a common nonsynonymous variant in *PM20D1*, p.Ile149Val (rs1891460), which is not in LD with the Parkinson’s Disease GWAS marker in this locus. Since this variant is not reported in ClinVar, has a low CADD score (1.01), and the statistical association is of borderline significance when correcting by the number of genes, it is possible that this association was found by chance. Replications in other cohorts will be required to determine whether this variant is associated with Parkinson’s Disease.

The associations described above were detected using different coverage thresholds for the detection of common and rare variants. For common variants, we used a threshold of coverage of ≥15X, which based on our previous studies (Rudakou *et al*., 2019; Krohn *et al*., 2019b; Rudakou *et al*., 2020; Krohn *et al*., 2020) is highly reliable for detection of known common variants. Here too, when comparing to available data from GWAS genotyping, we were able to confirm with 100% accuracy the presence of common variants in *MAPT*, *TMEM175* and *GPNMB* in samples with available data. The different thresholds of ≥ 30X and ≥ 50X are being used to increase our confidence that the variants and the results are real. The ≥ 50X threshold is highly reliable, yet also at ≥ 30X most detected variants are real. In the current study, the variants driving the associations in *FGF20* and *GCH1* has been fully confirmed with Sanger sequencing, further validating our results.

Our study has several limitations. The Parkinson’s disease patients and controls are not matched by sex and age, hence we adjusted for these variables in the statistical analysis. We found no significant difference between allele frequencies of young (<40 years old) and old controls, which allowed us to use all of the controls in our analyses. In addition, the coverage of some of the genes we analyzed was suboptimal (Supplementary Table 2), and we did not target intronic regions and other potentially regulatory regions outside the 5’ and 3’ UTRs, which may also affect gene function. Whole genome sequencing studies will be required to study these regions and overcome the coverage limitations. Another limitation of our study is the potential genetic heterogeneity, since cohorts that with both European or Ashkenazi Jewish patients and controls were included in the analyses. While we did include ethnicity as a covariate when needed, the Ashkenazi Jewish ancestry was a reported ancestry, not confirmed by GWAS data. Therefore, there could still be some residual effect of ethnicity, and our results should be confirmed in additional populations. Of note, when we removed the Ashkenazi Jewish individuals from the analysis, we yielded similar results, yet less significant due to the reduced power.

Our results, combined with previous studies, highlight the importance of performing fine mapping of GWAS loci to identify the variants and genes within each locus that drive the reported associations. Thus far, the causative genes within most Parkinson’s Disease GWAS loci are unknown, with the exception of *SNCA*, *GBA*, *LRRK2*, and likely *VPS13C*, *MAPT*, *TMEM175* and *GCH1*. The current study suggests that *SYT11, FGF20*, and potentially *PM20D1*, *BST1* and *GPNMB* should be considered as possible Parkinson’s Disease-related genes and targets for further research.

## Data Availability

Anonymized data is available upon request by any qualified investigator.

## Abbreviations

AJ: Ashkenazi Jewish
CADD: Combined Annotation Dependent Depletion
CI: confidence interval
CNS: central nervous system
DOPA: dihydroxyphenylalanine
DP: depth of coverage
eQTL: expression quantitative trait loci
GATK: Genome Analysis Toolkit
GnomAD: Genome Aggregation Database
GTEx: Genotype-tissue expression
GWAS: Genome-wide association study
hg19: human genome version 19
LD: linkage disequilibrium
LOF: loss-of-function
MAF: minor allele frequency
MIPs: molecular inversion probes
NS, Nonsyn: non-synonymous
OR: odds ratio
QC: quality control
REM: Rapid Eye Movement
SKAT-O: optimized sequence kernel association test
sQTL: splicing quantitative trait loci
UTR: untranslated region
yo: years of age

## Acknowledgements

We thank the participants for contributing to the study. GAR holds a Canada Research Chair in Genetics of the Nervous System and the Wilder Penfield Chair in Neurosciences. EAF is supported by a Foundation Grant from the Canadian Institutes of Health Research (FDN grant – 154301). ZGO is supported by the Fonds de recherche du Québec - Santé (FRQS) Chercheurs-boursiers award, in collaboration with Parkinson Quebec, and by the Young Investigator Award by Parkinson Canada. The access to part of the participants for this research has been made possible thanks to the Quebec Parkinson’s Network (http://rpq-qpn.ca/en/). We thank Daniel Rochefort, Helene Catoire, Sandra B. Laurent, Clotilde Degroot and Vessela Zaharieva for their assistance.

## Supplementary material

Supplementary Table 1: Detailed information on molecular inversion probes (MIPs)

Supplementary Table 2: Coverage statistics

Supplementary Table 3: All identified rare variants with 30x minimum depth of coverage

Supplementary Table 4: All identified rare variants with 50x minimum depth of coverage

Supplementary Table 5: All identified common variants

Supplementary Table 6: SKAT-O results for individual cohorts

Supplementary Table 7: FDR-corrected p values of all SKAT-O results at 30x and 50x minimal depth of coverage

>Supplementary Table 8: Results of SKAT-O analysis of early onset PD patients at coverage >=30X

>Supplementary Table 9: Results of SKAT-O analysis of early onset PD patients at coverage >=50X

>Supplementary Figure 1: The 3’UTR *GPNMB* variant (rs5850) is associated with reduced expression in numerous brain regions.

